# Real-world impact of a sepsis early detection model integrated into clinical workflow: a quasi-experimental study

**DOI:** 10.64898/2026.05.22.26353890

**Authors:** Yifan Zhang, Sarolta H. Trinh, Tom Phelan, Thomas F. Byrd, Roshan Tourani, Vipin Kumar, Pedro J. Caraballo, Genevieve B. Melton, Gyorgy J. Simon

## Abstract

**Background:** Sepsis is a life-threatening condition in which delayed recognition and treatment are associated with increased mortality. While predictive models such as Epic’s Early Detection of Sepsis Model (ESM) were developed to support early intervention, their real-world impact after integration into clinical workflows remains difficult to evaluate.

**Objectives:** To evaluate the real-world impact of ESM integrated into clinical workflow on clinical outcomes, antibiotic use, and harm-benefit tradeoffs.

**Methods:** We conducted a quasi-experimental study in a single healthcare system using encounter-level data from inpatient settings. Inpatient mortality, prolonged hospitalization, antibiotic use, and sepsis prevalence were compared between the pre-implementation period (3 June 2023 to 20 August 2024) and the online period (21 August 2024 to 26 December 2024) when the model became visible to clinicians. We also applied a counterfactual framework using models trained on pre-implementation data to estimate expected outcomes without ESM and to quantify harms related to overtreatment and delayed treatment.

**Results:** Among 101,138 encounters, 86,884 occurred during the pre-implementation period and 14,254 during the online period. In unadjusted analyses, the online period had lower inpatient mortality, prolonged hospitalization, antibiotic use, and sepsis prevalence (all p≤0.002). In the counterfactual analyses, observed outcomes were lower than expected without ESM for mortality (1.21% vs 1.82%; p<0.001), prolonged hospitalization (5.56% vs 7.95%; p<0.001), and antibiotic use (43.52% vs 47.04%; p<0.001). False positive harm (37.72% vs 41.68%; p<0.001) was also lower than expected.

**Conclusions:** Integration of ESM into clinical workflow was associated with improved patient outcomes, reduced antibiotic use, and decreased harm from overtreatment, without evidence of increased harm from delayed treatment, supporting a positive net clinical benefit and the safety and effectiveness of ESM under Software as a Medical Device principles.

## 1. Background and Significance

Sepsis remains a major cause of preventable death, contributing to an estimated 11.0 million deaths globally and persistently high in-hospital mortality in the United States.^1,2^ Timely recognition and treatment of sepsis are essential, as delayed antibiotic initiation is associated with increased mortality.^3–7^ Because early sepsis recognition is both time-critical and clinically challenging due to the heterogeneity of etiology and presentation, electronic health record (EHR)-based machine learning models to support its early detection have been developed and deployed widely into clinical care.^8^

While there have been over a dozen evaluations of deployed sepsis detection tools, these manuscripts fail to evaluate the effects of the model and the decision support rigorously around workflow, model effects, and decision tradeoffs. Several studies rely on simple pre-post comparison, which can be biased when the population or severity shifts over time.^9–13^ Some have utilized a quasi-experimental evaluation, which do not isolate model-attributable effects.^14^ Beyond statistical problems, the clinical value of these models remains unclear, since most evaluations do not consider the tradeoffs between risks of harms and benefits from true, false and missed alerts.^15–17^

Although a randomized controlled trial (RCT) would provide the strongest causal evidence for the impact of a deployed model, RCTs are often impractical if not infeasible due to the cost and lack of feasibility. Once the model is implemented in the routine workflow, randomization and blinding are difficult because alerts are visible to clinicians and may influence care processes across patients and teams.^18^

## 2. Objectives

In this study, we evaluated the impact of implementing Epic’s Early Detection of Sepsis Model (ESM) Version 2 on patient outcomes. ESM alerts care teams of patients who may be in the early stages of sepsis based on the Sepsis-3 definition in the inpatient setting.^19,20^ We use a quasi-experimental design across a single integrated healthcare delivery system, including unadjusted comparisons, subpopulation analyses and counterfactual estimation to quantify ESM-attributable effects on clinical outcomes, antibiotic use, and harm-benefit tradeoffs related to false-positive and false-negative alerting.

## 3. Methods

### 3.1. Setting

This study was conducted at a single academic healthcare delivery system in the Midwest and included adult encounters in the inpatient setting across 9 hospitals. During the entire study period, there were 101,138 total encounters among 77,651 unique patients. Sepsis encounters with qualifying antibiotic administration before admission time and encounters without model predictions in the online period were excluded.

### 3.2. Study Design

A quasi-experimental pre-post evaluation of ESM was performed at the encounter level to assess changes in harms, outcomes and antibiotic use. We defined the **pre-acquisition** period from 3 June 2023 to 24 June 2024, covering a one-year period before the model was implemented; the **background validation** period from 25 June 2024 to 20 August 2024 during which the ESM was running in the background and predictions were not revealed to the care team; and the **online** period from 21 August 2024 to 26 December 2024 (4 months), when the ESM was integrated into the clinical workflow and predictions were visible. Since the *pre-acquisition* and *background validation* periods had no significant differences, they were merged into a single 14-month **pre-implementation** period (3 June 2023 to 20 August 2024).

### 3.3. ESM

ESM Version 2 is a locally trainable gradient-boosted tree (LightGBM) model based on 66 EHR predictor variables, including demographics, diagnoses, vital signs, laboratory results, flowsheets, nursing assessments, medications, procedures and utilization. The model updates the score every 30 minutes and predicts the risk of Sepsis-3 onset within the next 8 hours.^20^

### 3.4. Sepsis and related definitions

We follow Epic’s definition of **sepsis**, defined as clinically treated sepsis, based on the Sepsis-3 criteria with antibiotic use.^19^ Sepsis-3 requires suspected infection, identified by a qualifying body fluid culture collection and antibiotic administration (except single-dose antibiotics or those administered in the operating room), and organ dysfunction, defined by a qualifying SOFA increase or ≥ 2 points.^20^ In routine care, some encounters might meet criteria and have antibiotics started briefly but then discontinued, which may indicate that ultimately clinicians did not treat the patients as septic.^21^ Accordingly, we classified encounters as septic only if they met Sepsis-3 criteria and received antibiotics *that were not discontinued*.

Sepsis **onset** is defined as the earliest time (SIRS2 time) patients meet the Systemic Inflammatory Response Syndrome (SIRS2) criteria within an encounter.^22^ Even though SIRS2 is highly sensitive but not specific to sepsis, it provides an early and consistent sign of deterioration. We used it as the time to measure antibiotic timing and define treatment delay across encounters. For encounters with antibiotics before any observed SIRS2 time, we used admission time.

### 3.5. Primary outcomes

The primary outcomes of our evaluation were harms related to unnecessary treatment (false positive; FP), harms related to the lack of treatment (false negative; FN), clinical outcomes (inpatient mortality and prolonged ≥ 10 days of hospitalization), and antibiotic use.^23^

False positive harm (FPH) was defined as non-septic patients receiving antibiotics and was operationalized as antibiotic discontinuation (<3 days of antibiotic course).^6,21^ False negative harm (FNH), in which septic patients have delayed antibiotics, is defined as antibiotics being given more than 6 hours after SIRS2 time, as suggested in a prior study.^7^ When patients do not present with SIRS2 before antibiotic administration, we consider the intervention timely and not counted as a false negative.

### 3.6. Treatment Definition

Antibiotic treatment timing was defined as the earliest between the antibiotic administration time and the medication order time. Encounters with antibiotics before admission were excluded. When SIRS2 existed, antibiotic timing was assessed relative to the earliest SIRS2 time; otherwise, timing was assessed relative to admission time. For septic encounters, we defined timely treatment as antibiotics given within 6 hours of SIRS2 time and delayed treatment as antibiotics given more than 6 hours after SIRS2 time. As previously mentioned, we used antibiotic discontinuation to distinguish between sepsis and non-sepsis encounters by checking if there was a subsequent antibiotic order after 3 days.

### 3.7. Statistical analysis

#### 3.7.1. Differences between pre-implementation and online cohort characteristics

Cohort differences were assessed using the Kruskal-Wallis rank sum test for the continuous variables and Pearson’s chi-squared test for the categorical variables. Continuous variables are reported as median [IQR] and categorical variables as count (%). All hypothesis tests used a 95% significance level.

#### 3.7.2. Raw effect of ESM

The unadjusted effect associated with ESM implementation was evaluated by comparing encounter-level rates of primary outcomes, including inpatient mortality, prolonged hospitalization, antibiotic use, and sepsis prevalence between the pre-implementation and the online period. For each comparison, we summarized the effect size using the relative percent change from the pre-implementation period to the online period and used a two-proportion z-test.

#### 3.7.3. Subgroup analysis

To further assess whether changes in sepsis diagnosis in the online period were related to the shifts in antibiotic use, we stratified encounters by sepsis status and antibiotic treatment decision into four workflow outcome groups. The subgroup analyses were conducted by comparing proportions of clinical outcomes between the pre-implementation and online periods within FP and FN groups, as well as among septic patients with timely treatment (true positive; TP) and non-septic patients without treatment (true negative; TN). Since the TN group is heterogeneous in terms of outcome risks, we further analyzed those TN patients who presented with SIRS2. Differences were evaluated using two-proportion Z-tests.

#### 3.7.4. Effect of ESM adjusted for severity

Since our study uses a pre-post comparison, factors other than ESM may have influenced the outcomes, including differences in patient severity, sepsis prevalence, and treatment probability over time.^24^ To reduce these biases, we specified a causal structure and the corresponding estimators using predictive models trained on pre-implementation data to estimate primary outcomes in the counterfactual online period scenario without ESM. The counterfactual estimates were compared with the observed rate to quantify the effect of ESM, and the two-proportion Z-test was applied.

##### 3.7.4.1. Data acquisition

We constructed a retrospective EHR-derived dataset for the pre-implementation and the online periods. The dataset contained encounter-level clinical outcomes, patient characteristics (age and legal sex), diagnosis of urinary-tract infection, sepsis status, and treatment indicator. For each sampled timestamp, we retrieved the most recent laboratory and vital signs measurements recorded within the preceding 72 hours. All variables are selected from the ESM documentation. In total, 23 variables were available in the EHR and were included.

##### 3.7.4.2. Sampling

To convert variable-length longitudinal data into fixed-length sequences for counterfactual modeling, we used Epic’s time-slicing approach,^20^ with a necessary modification. For positive encounters (sepsis cases), sampling was performed at 30-minute intervals over the 8 hours preceding the antibiotic administration, yielding 15 fixed samples (excluding the time of the antibiotic administration); and for negative encounters (no sepsis), 15 random samples were taken from the entire time series. With antibiotics being part of the sepsis definition, all septic patients generally received antibiotics. The modification concerns the number of samples. We found that with fixed 15 samples, outcome leakage would occur. The reason is differential imputation: while we almost always have 15 samples for negative patients, for positive patients, who mostly received antibiotics well within 8 hours, we would perform so much imputation that it would distort their predictor distributions. To avoid this, we reduced the number of samples to 6.

##### 3.7.4.3. Imputation

At each sampled time point, laboratory results and vital signs were carried forward from the most recent available value within the prior 72 hours. If no value was available, we used mode (for categorical variables) and median (for continuous variables) imputation based on the pre-implementation data.

##### 3.7.4.4. Modeling

To estimate the counterfactual outcomes that would have occurred during the online period without the ESM model, we specify a causal structure shown in Fig. 1. In the presence of ESM predictions *M*, patient characteristics *X* causes predictions *M*, sepsis status *Y*, treatment decision *T* and clinical outcomes *Z*. *M* provides an additional path to *T*, which can affect downstream outcomes. Because our sepsis definition incorporates antibiotic use (treatment *T*), *T* also causally affects *Y*. In the counterfactual setting without ESM, causal structure follows that of the pre-implementation period, with the pathway through *M* removed.

**Figure 1.**
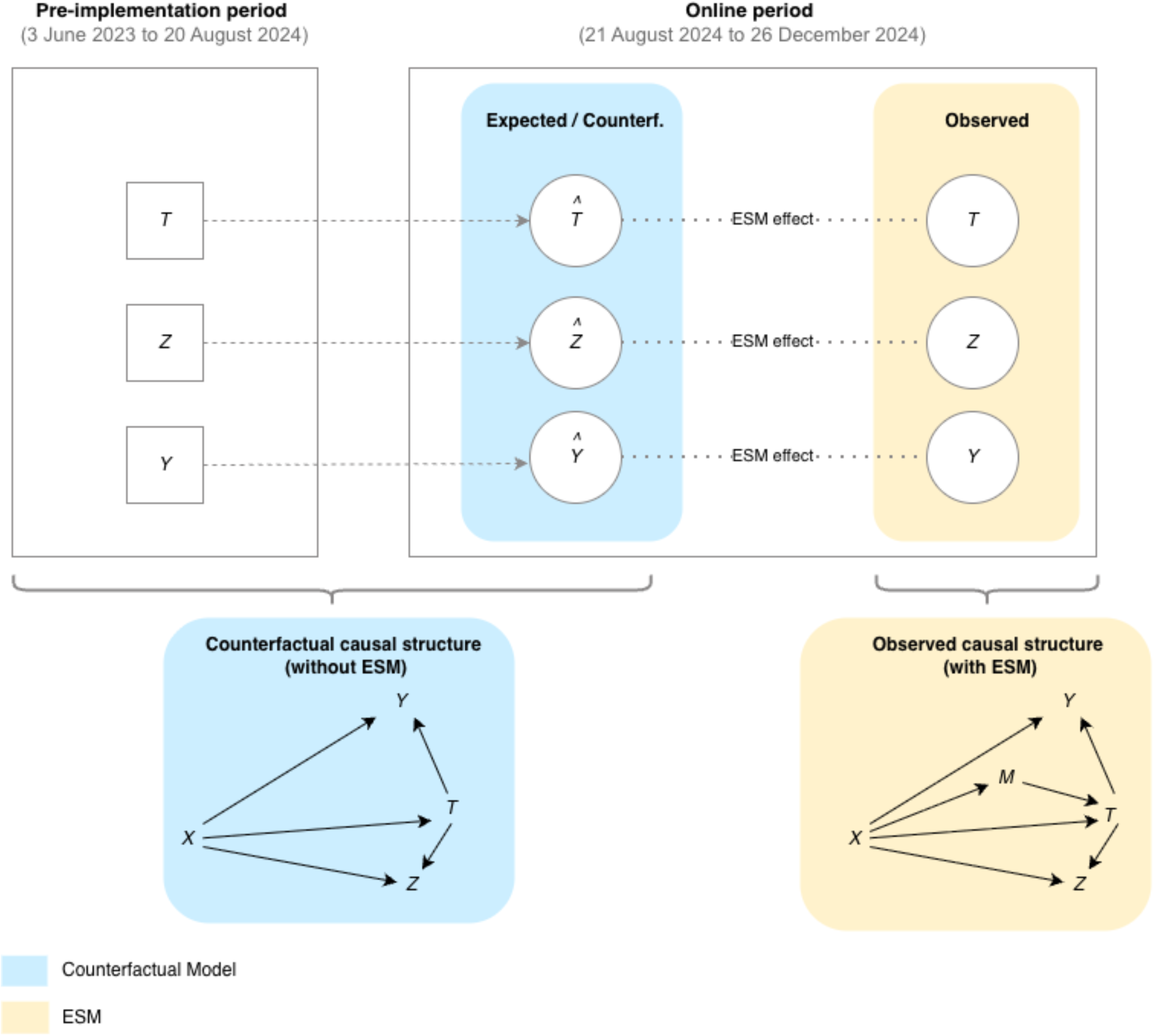
Summary of the counterfactual models and the underlying causal structure for estimating ESM-attributable effects. Squares represent the counterfactual models for predicting treatment *T*, outcome *Z*, and sepsis *Y* trained on patient characteristics *X* from the pre-implementation period. These were applied to estimate counterfactual outcomes during the online period and were compared with the observed outcomes obtained using the ESM (*M*). Blue indicates the counterfactual model framework, and beige indicates the observed ESM-based framework.

Based on this structure, we estimated counterfactual rates in the online period using three predictive models trained on the pre-implementation period: (1) a treatment model 𝑃(𝑇|𝑋), (2) an outcome model 𝑃(𝑍|𝑇, 𝑋), and (3) a sepsis model 𝑃(𝑌|𝑇, 𝑋). The ESM-attributable effects were then quantified as the difference between the observed values in the online period and the corresponding counterfactual expected values. Using the treatment and outcome models, the counterfactual expected outcome rate was computed from the encounter-level probabilities as

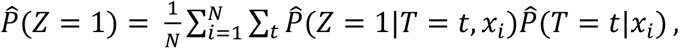

where *t* = 1 indicates antibiotic administration and *t* = 0 indicates no timely treatment.

We also quantified harms related to the treatment decisions and sepsis status within the same structure. FPH and FNH were defined as 𝐹𝑃𝐻 = {𝑌 = 0, 𝑇 = 1} and 𝐹𝑁𝐻 = {𝑌 = 1, 𝑇 = 0}. These probabilities in the online period were estimated as follows:

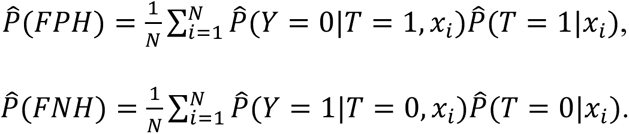

All models were fit using logistic regression with pre-implementation encounters only, and model performance was evaluated on a hold-out validation set to assess overfitting. To avoid patient-level information leakage, we split the pre-implementation dataset by patient identifier, assigning 80% of patients to training and 20% to validation.

##### 3.7.4.5. Generating the results

For each primary outcome, we summarized ESM-associated changes in the online period using both the observed rate (“actual”) and the counterfactual rate (“expected”) obtained by applying the pre-implementation trained models to online encounters. The adjusted effect was expressed as the absolute difference between the expected and observed rates and the relative percent difference, defined as |𝐸𝑥𝑝𝑒𝑐𝑡𝑒𝑑 − 𝐴𝑐𝑡𝑢𝑎𝑙|/𝐸𝑥𝑝𝑒𝑐𝑡𝑒𝑑 × 100%. Observed and expected rates were compared using two-proportion Z-tests.

## 4. Results

### 4.1. Study population

In the final study population, 86,884 encounters from 64,818 patients were included in the pre-implementation period, and 14,254 encounters from 12,833 patients in the online period. As shown in Table 1, at the patient level, the online period had a lower mortality rate (2.5% vs 1.3%; p<0.001) and a lower proportion of prolonged hospitalization (9.9% vs 5.9%; p<0.001). Several baseline differences were observed, including age and historical urinary tract infection, as well as several laboratory and vital sign variables.

**Table 1.**
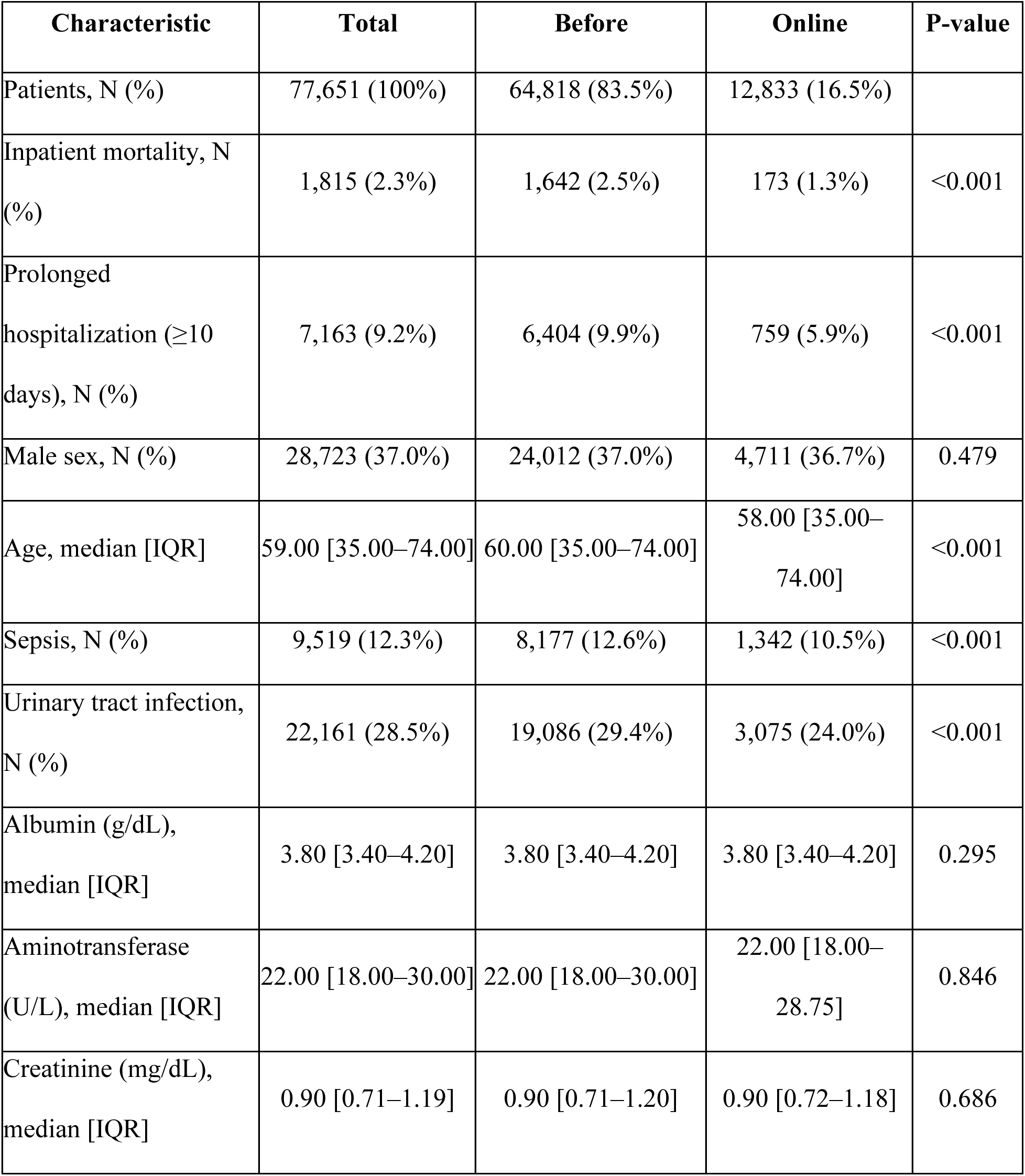
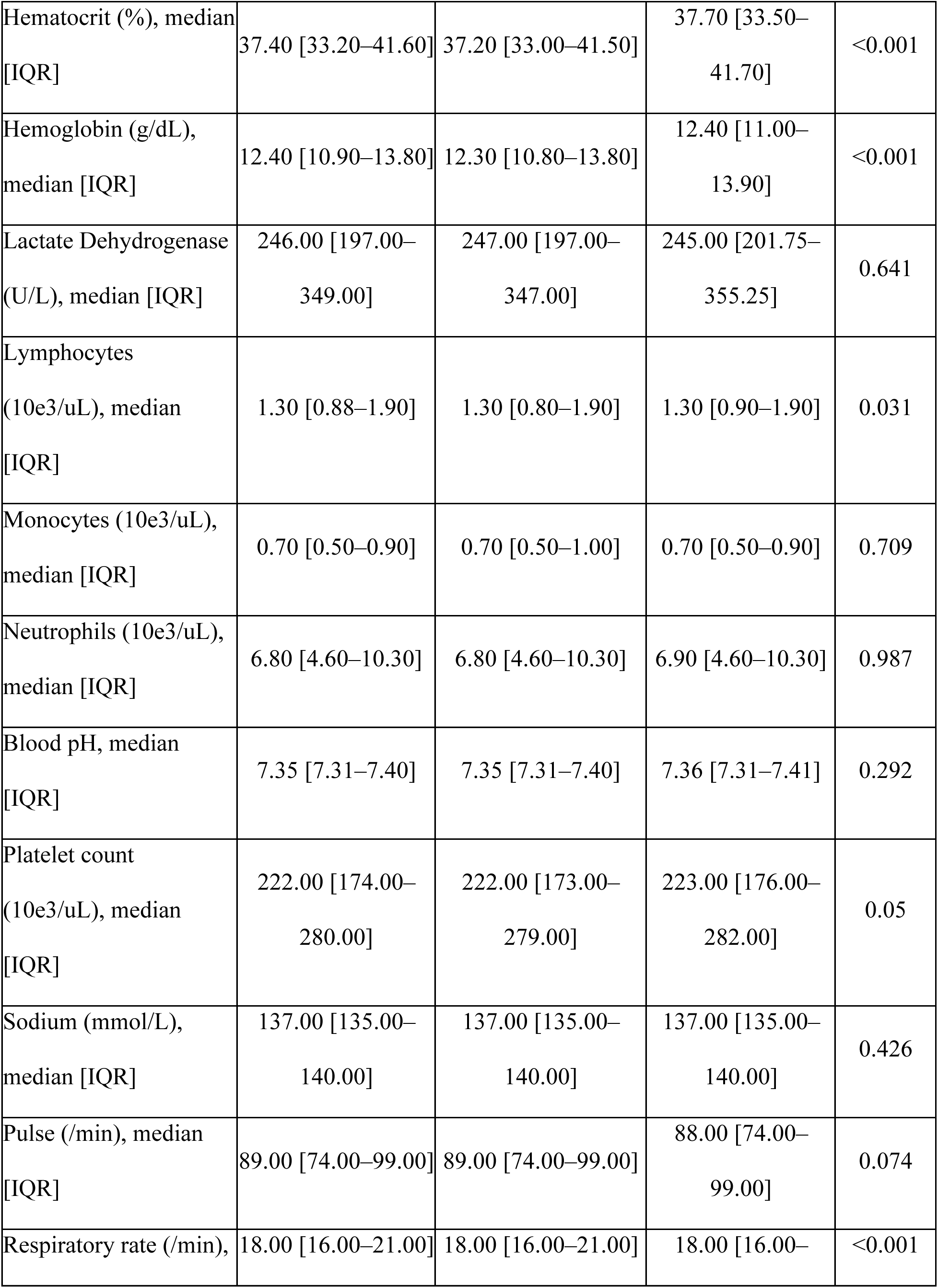
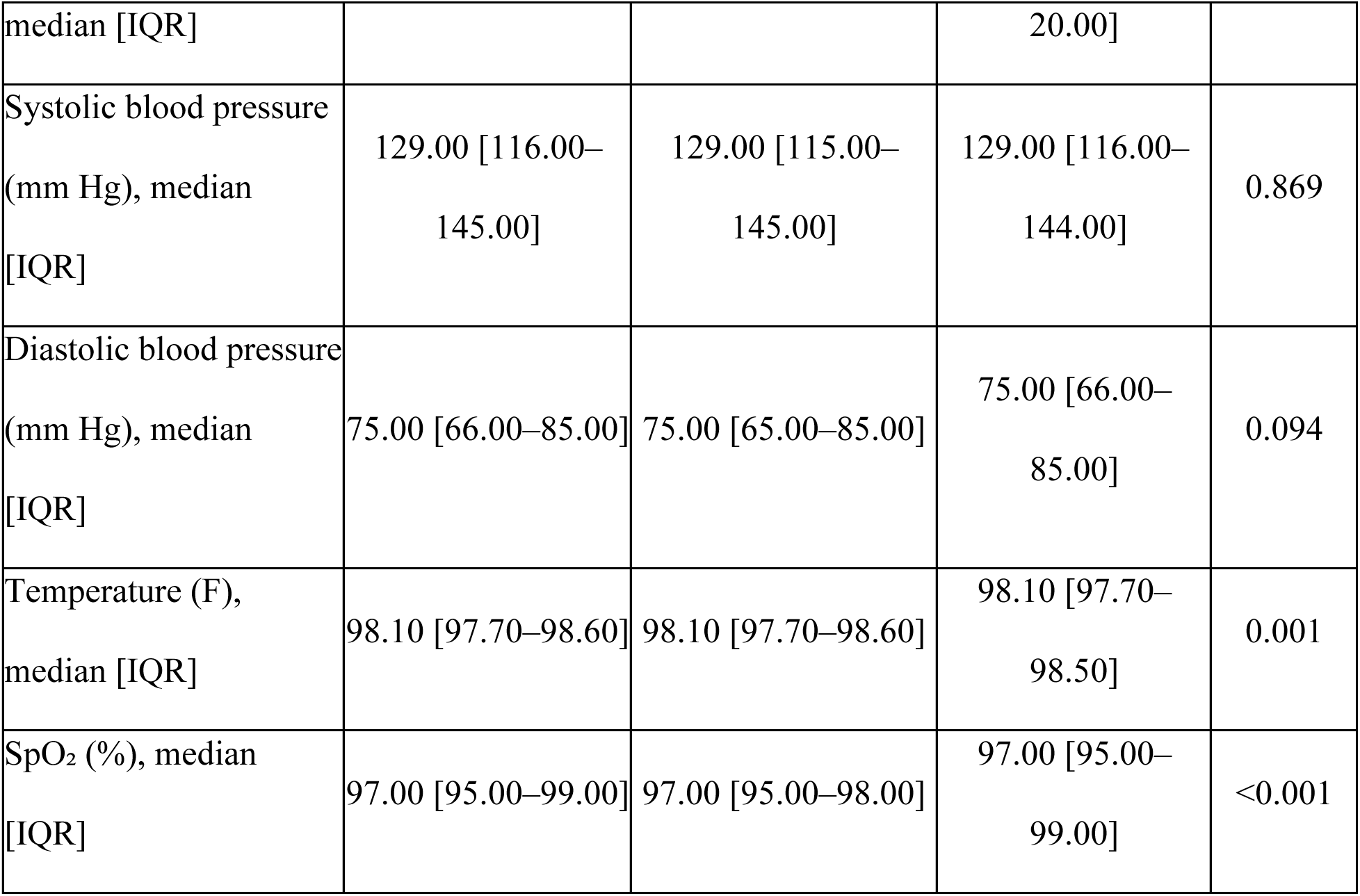
Demographics and baseline characteristics of patients in the pre-implementation and online period.

### 4.2. Unadjusted comparison

Encounter-level main observed changes between pre-implementation and online periods are summarized in Table 2. Inpatient mortality decreased from 1.89% in the pre-implementation period to 1.21% in the online period, corresponding to a 35.98% relative decrease (p<0.001). The probability of prolonged hospitalization decreased from 8.55% to 5.56%, a 34.97% relative decrease (p<0.001). Antibiotic use also decreased from 49.58% to 43.52% (12.22% relative reduction; p<0.001). The observed sepsis proportion decreased from 10.50% to 9.67% (7.90% relative reduction; p=0.002).

**Table 2.** The main observed differences between pre-implementation and online periods.

|  | **Pre-implementation** | **Online** | **Relative Diff** | **P-value** |
| --- | --- | --- | --- | --- |
| N | 86,884 | 14,254 |  |  |
| Mortality | 1.89% | 1.21% | 35.98% | <0.001 |
| Prolonged Hospitalization | 8.55% | 5.56% | 34.97% | <0.001 |
| Antibiotics Use | 49.58% | 43.52% | 12.22% | <0.001 |
| Sepsis | 10.50% | 9.67% | 7.90% | 0.002 |

### 4.3. Subgroup analysis

The comparisons within subgroups on the clinical outcomes between pre-implementation and online period are shown in Table 3. Mortality decreased significantly in the online period among FP (1.74% vs 0.84%; p<0.001), TN (0.87% vs 0.51%; p=0.001), and TN with SIRS2 (1.35% vs 0.84%; p=0.012). Prolonged hospitalization decreased significantly in all subgroups except TN without SIRS2. Mortality differences were not significant for the remaining subgroups.

**Table 3.** Comparisons between pre-implementation and online periods on observed clinical outcomes within each subgroup.

|  | Pre-implementation (%) |  | Online (%) |  | P-value |  |
| --- | --- | --- | --- | --- | --- | --- |
|  | Mortality | Prolonged Hospitalization | Mortality | Prolonged Hospitalization | Mortality | Prolonged Hospitalization |
| TP | 6.05 | 15.65 | 5.45 | 12.83 | 0.496 | 0.037 |
| FP | 1.74 | 10.25 | 0.84 | 6.73 | <0.001 | <0.001 |
| FN | 8.27 | 37.34 | 8.15 | 30.80 | 0.922 | 0.003 |
| TN | 0.87 | 3.25 | 0.51 | 2.05 | 0.001 | <0.001 |
| TN w/<br>SIRS2 | 1.35 | 5.35 | 0.84 | 3.39 | 0.012 | <0.001 |
| TN w/o<br>SIRS2 | 0.36 | 1.06 | 0.20 | 0.84 | 0.123 | 0.220 |

### 4.4. Counterfactual analysis of the effect of ESM

After describing the unadjusted differences in primary outcomes, we evaluated whether these changes were consistent under a counterfactual scenario in which ESM was not implemented (Table 4). The observed inpatient mortality rate in the online period was 1.21%, while the expected counterfactual mortality was 1.82%, corresponding to a 33.33% relative decrease (p<0.001). Similarly, the prolonged hospitalization occurred less than expected without the ESM (5.56% vs 7.95%), a 30.10% relative reduction (p<0.001). Antibiotic use was lower than expected (43.52% vs 47.04%), a 7.50% relative reduction (p<0.001).

**Table 4.** Observed outcomes in pre-implementation and online period, the expected counterfactual values based on the causal structure.

|  | Pre-implementation<br>n (%) | Online (%) |  | Relative<br>Diff (%) | P-value |
| --- | --- | --- | --- | --- | --- |
|  |  | Actual | Expected |  |  |
| Mortality | 1.89 | 1.21 | 1.82 | 33.33 | <0.001 |
| Prolonged<br>Hospitalization | 8.55 | 5.56 | 7.95 | 30.10 | <0.001 |
| Abx Use | 49.58 | 43.52 | 47.04 | 7.50 | <0.001 |
| Sepsis | 10.50 | 9.67 | 9.69 | 0.26 | 0.944 |
| FPH | 43.47 | 37.72 | 41.68 | 9.50 | <0.001 |
| FNH | 4.40 | 3.87 | 4.31 | 10.19 | 0.061 |
| TP | 6.11 | 5.79 | 5.36 | 8.02 | 0.112 |
| TN | 46.03 | 52.61 | 48.64 | 8.15 | <0.001 |

For harm measures, the FPH rate was lower (37.72% vs 41.68%), corresponding to a 9.50% relative reduction (p<0.001). For completeness, we also included the TP and TN groups, with the TN being higher than expected counterfactual (52.61% vs 48.64%), corresponding to a 8.15% relative increase (p <0.001).

## 5. Discussion

Our multi-pronged evaluation of ESM implementation in clinical workflow within an integrated healthcare delivery system showed improved clinical outcomes. Compared with the pre-implementation period, the online period had lower inpatient mortality, fewer prolonged hospitalizations, and lower antibiotic use, along with a decrease in sepsis prevalence. To address potential bias from the pre-post comparison, we compared observed online outcomes with counterfactual expectations without ESM and found lower mortality, prolonged hospitalization and antibiotic use than expected. Harm-benefit tradeoffs were also assessed using harm metrics. Relative to the counterfactual expectation, FPH and FNH were lower, while TP and TN were higher. Together, these findings support an association between ESM availability and improved outcomes alongside more appropriate antibiotic use.

With ESM being a detection rather than a prevention tool, no changes in sepsis prevalence would be expected between the pre- and post-implementation periods, yet a significant decrease was observed. We considered several additional potential explanations. First, it is a seasonal variation, as sepsis rates are known to peak in winter and be lowest during summer.^25^ Second, it may reflect an ESM-related effect, with earlier recognition and treatment preventing progression to overt sepsis. Third, the change could be driven by the sepsis definition used in this study, which incorporated antibiotic use, so the lower sepsis prevalence may merely reflect reduced full-course antibiotic use. With no adverse impact on the primary outcomes, it is conceivable that sepsis was overdiagnosed in the pre-implementation period. Fourth, differences in hospital population severity may have contributed to the observed decrease in sepsis prevalence, as predicted mortality risk differed between periods at both admission and antibiotic administration (Fig. S1). However, this difference was mitigated in counterfactual analysis, emphasizing that the unadjusted decrease should be interpreted cautiously. Finally, we cannot exclude another concurrent factor, such as increased sepsis awareness due to ESM-related training.

If the reduced sepsis diagnosis were primarily driven by reduced antibiotic use rather than reduced prevalence, a concern of underdiagnosis in the post-implementation period arises. In that case, some septic encounters may have been missed initially and received delayed antibiotics, whereas others may have been misclassified as non-septic because antibiotics were not given. Worse outcomes would be expected in the FN (corresponds to delayed treatment) and TN (corresponds to unrecognized sepsis) groups, especially in TN with SIRS2 group, who are at higher outcome risk. We did not observe this pattern in the subgroup analysis, which argues against underdiagnosis as the primary explanation for the lower sepsis prevalence.

Despite the growing number of sepsis prediction models with strong predictive performance, their real-world clinical impact remains unclear. Few have been deployed in clinical workflow and even fewer have been evaluated after implementation using standardized approaches. Assessing the impact of models implemented in the workflow is challenging because blinding and randomization are often infeasible or impractical once the model is integrated into routine care, which makes rigorous analytical methods necessary and all the more critically important. In this study, we presented a disciplined approach that combines quasi-experimental design with counterfactual estimation to isolate model-attribute effects.

Additionally, to our knowledge, this is the first study that focuses on the clinical value of a deployed sepsis model, and the first to quantify its impact as seen through a risk management lens by considering harm-benefit tradeoffs.

Prior deployment studies evaluating the effect of a sepsis detection model in a pre-post design exist. Topiwala et al. focus on alert performance and timeliness, while Giannini et al. and Hsu et al. rely largely on unadjusted before-after comparison in processes, utilization, and mixed mortality effects.^10,13,15^ In contrast, our study specified a causal structure and regression-based counterfactual estimation to reduce bias in raw pre-post comparisons. Even stronger designs, such as the randomized trial by Shimabukuro et al. and the Bayesian causal impact analysis by Boussina et al., mainly focused on mortality or length of stay without formally quantifying the clinical tradeoffs of acting on alerts.^14, 17^ Moreover, we combine the FPH and FNH to quantify net clinical value overall and within subgroups, enabling a more detailed interpretation of effects across multiple perspectives.

The two key principles underlying FDA’s clinical AI regulation (Software as Medical Device, SaMD) are (1) harms from the use of the model must be acceptably low [safety] and (2) benefits from using the model must outweigh any potential harms [efficiency]. In the pre-implementation period, the health system had 9.88 FPs (patients receiving less than full course antibiotics) per FN (patient with delayed antibiotic treatment), yielding a FN:FP tradeoff of 1/9.88 = 0.101. Assuming that this tradeoff and both harms were acceptable in the pre-implementation period, the net benefit^26,27^ of ESM can be computed as the increase in TPs (from the counterfactual 5.36 per 100 patients to the observed 5.79) discounted by the increase in FPs (from the counterfactual 41.68 to the observed 37.72 per 100 patients, which actually is a *decrease*) multiplied by the weighing factors, giving a net benefit of 𝑇𝑃 − 𝑤 ∗ 𝐹𝑃 = 0.43 − 0.101 ∗ (37.72 − 41.68) = 0.83. Discounted for FPs, ESM correctly identified an additional 0.83 septic patient per 100 patients. Without the ESM, the discounted TPs would have been 1.15 patients per 100, so ESM increased this by approximately 72%. Notably, FPs decreased rather than increased with increasing TPs, meaning the “discount” for FPs became a “reward”. The reduction in FPH, without evidence of increased FNH, and the positive net benefit support the safety and effectiveness of ESM.

Our study has several limitations. First, because this is a non-randomized evaluation, even though we used a prespecified causal structure to reduce bias, residual latent confoundings may remain from concurrent practice changes that are not fully captured by counterfactual modeling, such as clinical awareness or other models running during the same period. Second, the evaluation was conducted in a single health system, so effect sizes may limit generalizability to settings with different workflows, patient populations, implementation strategies, or alert governance. Finally, our outcomes depend on how sepsis was identified by the Sepsis-3 definition, incorporating antibiotic use, so misclassification is possible, including patients with noninfectious syndromes with similar symptoms.

## 6. Conclusions

Implementation of ESM in clinical workflow was associated with improved clinical outcomes, reduced inappropriate antibiotic use, reduced harm from overtreatment, and no evidence of increased harm from delayed treatment in a real-world setting. More importantly, our study provides a strong proof of concept for a more rigorous and disciplined approach to post-deployment evaluation of clinical prediction models by combining counterfactual estimation with harm-benefit assessment. This framework may support more credible evaluation of the safety, effectiveness, and clinical value of future AI tools in practice.

## Data Availability

The individual-level data analyzed in this study are not publicly available because they contain protected health information derived from electronic health records. Aggregate results are reported in the manuscript and supplementary materials.

## Clinical Relevance Statement

Implementation of ESM into clinical workflow was associated with improved patient outcomes, including lower inpatient mortality, fewer prolonged hospitalizations, and reduced antibiotic use. This study also provides a practical approach for evaluating deployed clinical AI in real-world settings when randomization is not feasible. The findings support the safety and effectiveness of the model to improve clinical outcomes while reducing inappropriate treatment.

## Multiple Choice Questions

1. When evaluating the real-world impact of a prediction model after it is integrated into clinical workflow, which method in this study was used to reduce bias from simple pre-post comparison?

A. Clinician survey
B. Patient randomization
C. Counterfactual estimation
D. Case-control matching

Answer: C. Simple pre-post comparisons can be biased, in the case of this study, because patient severity, sepsis prevalence and treatment probability change over time. A counterfactual estimation addresses this by comparing observed outcomes with expected outcomes predicted from pre-implementation data.

2. Which metric combines true positives and false positives using a harm-based weighting factor to evaluate overall clinical value?

A. Recall
B. Net benefit
C. Specificity
D. F1-score

Answer: B. Net benefit incorporates both correct positive decisions and the penalty for false positives after applying a weighting factor that reflects the acceptable tradeoff between harms and benefits.

## Acknowledgments

This work was supported by the Minnesota Partnership for Biotechnology and Medical Genomics Grant (project number: 00110451), the National Center for Advancing Translational Sciences of the National Institutes of Health (NIH) (grant number: UM1TR004405), and the University of Minnesota Center for Learning Health System Sciences, a collaboration between the Medical School, the School of Public Health and its training program, the Minnesota-LHS Scholars Program funded by the Agency for Healthcare Quality and Research (AHRQ) and the Patient-Centered Outcomes Research Institute (PCORI) (grant number P30HS02744, TB). The funders had no role in the study design, data collection, analysis, decision to publish, or manuscript preparation. The content is solely the responsibility of the authors and does not represent the official views of the University of Minnesota, Mayo Clinic, NIH, AHRQ, or PCORI.

## Conflict of Interest

The authors declare that they have no conflicts of interest in the research.

## Human Subjects Protections

The study was approved by the institutional review board under protocol STUDY00019710.

## Notes

### Competing Interest Statement

The authors have declared no competing interest.

### Author Declarations

The Institutional Review Board of the University of Minnesota gave ethical approval for this work.

