## Supplementary material for "Real-world impact of a sepsis early detection model integrated into clinical workflow: a quasi-experimental study": Fig. S1

**Figure S1.** Predicted mortality risk at admission and antibiotic administration between pre-implementation and online periods. Predicted mortality risk was estimated using a logistic regression mortality model trained on pre-implementation encounters, with predictors measured at admission and antibiotic administration, and then applied to online-period encounters.

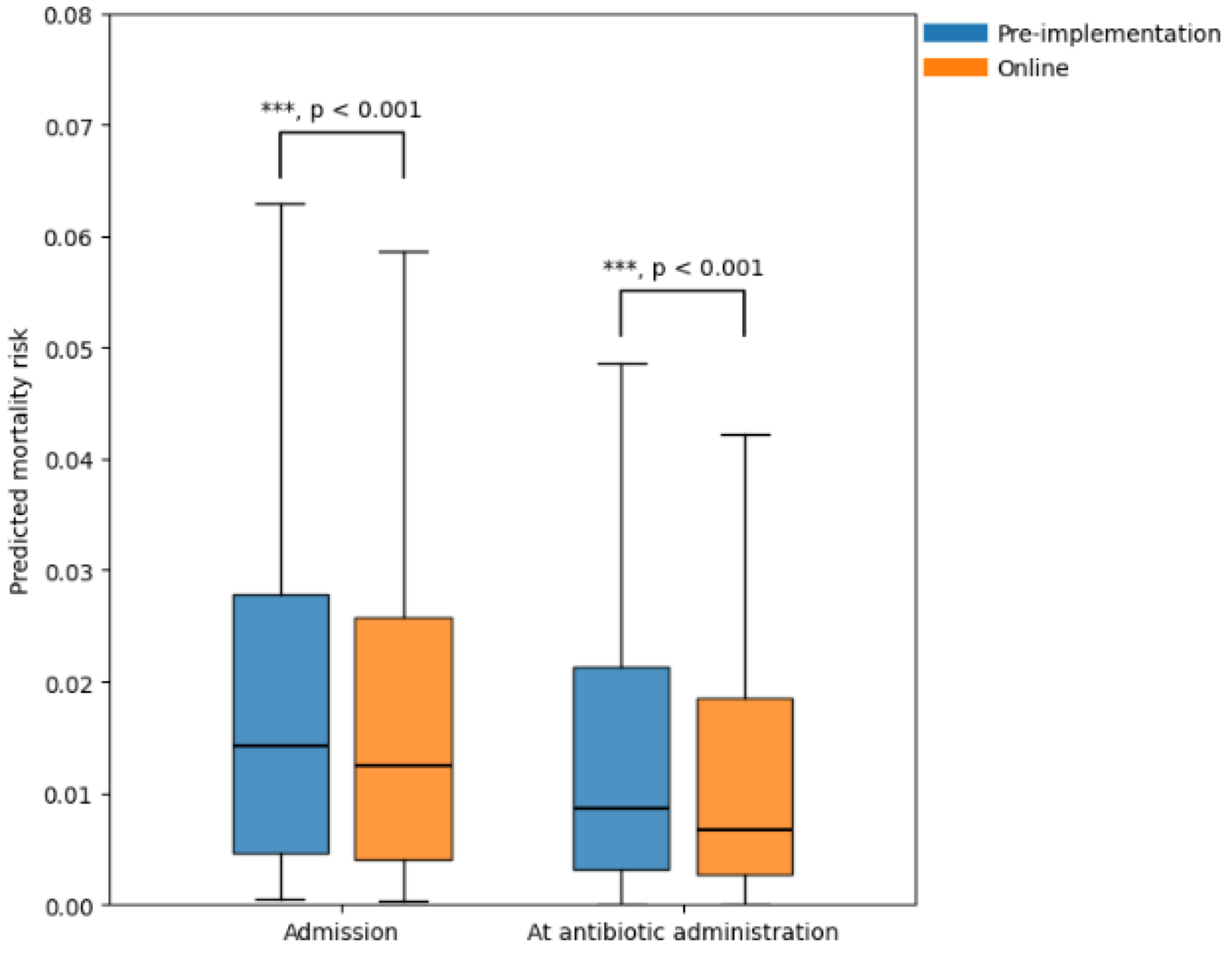
